# Clonal Transmission of Emerging Novel *Plasmodium falciparum* Kelch13 Mutations and Increasing Complexity of Infection in Libreville, Gabon, 2021-2023

**DOI:** 10.1101/2025.08.07.25333205

**Authors:** Jacques Mari Ndong Ngomo, Denise Patricia Mawili-Mboumba, Tobbi Ndong Mouity, Alec Leonetti, Dimitri Moussavou Mabika, J’ose Coell Mihindou, Noé Patrick Mbondoukwé, Jonathan J. Juliano, Jeffrey A. Bailey, Marielle Karine Bouyou-Akotet, Karamoko Niaré

**Author notes:** corresponding authors: Karamoko Niare, PhD, Department of Pathology and Laboratory Medicine, Brown University, Providence, RI, USA, Jacques Mari Ndong Ngomo, Département des Sciences fondamentales, service de Parasitologie-Mycologie et de Médecine Tropicale, Université des Sciences de la Santé. equal senior author contribution (Jeffrey A. Bailey, Marielle Karine Bouyou-Akotet, Karamoko Niaré).

## Abstract

Artemisinin partial resistance (ART-R) in *Plasmodium falciparum*, due to mutations in the Kelch13 (K13) propeller domain, is spreading across Africa. However, data from Central Africa remain sparse. This study performed molecular surveillance in a peri-urban sentinel site in Libreville, Gabon, from 2021 to 2023 to assess emerging resistance markers and parasite population dynamics. Febrile patients with confirmed *P. falciparum* infection were enrolled at the Melen sentinel site. Dried blood spots were collected and isolated DNA sequenced using molecular inversion probes (MIPs) targeting drug resistance genes and genome-wide SNPs. We assessed the prevalence of mutations in K13, DHFR, DHPS, CRT, and MDR1. Complexity of infection (COI) and identity-by-descent (IBD) were used to evaluate transmission intensity and parasite relatedness, respectively. Among 468 genotyped samples, no validated or candidate K13 mutations were detected. However, 21 carried K13 mutations of unknown significance, including E433D (n=14), Q613H (n=5), V520I, and V637I. Interestingly, E433D prevalence rose from 0.7% in 2022 to 5.5% in 2023. Parasites with E433D or Q613H showed significantly higher IBD than wild-type (P<0.001) and chains of clonal transmission. Recent DHFR and DHPS mutations associated with higher-level sulfadoxine-pyrimethamine resistance were at low prevalence. MDR1 Y184F increased from 52.6% to 68.8%, while CRT K76T remained rare. IBD estimates support clonal transmission of parasites carrying emerging K13 mutations, particularly E433D and Q613H. In parallel, COI estimates increased over time, suggesting intensifying malaria transmission, potentially with a seasonal component. These findings highlight the need for expanded genomic surveillance and functional validation of these novel mutations to inform malaria control strategies in Gabon and Central Africa.

## INTRODUCTION

Malaria continues to pose a significant global health challenge, particularly in sub-Saharan Africa, where it causes high rates of illness and death (1). Despite decades of intervention efforts, progress has been inconsistent, with only minimal declines in malaria burden across the region in recent years. Several factors threaten the effectiveness of malaria control, including resistance to drugs, diagnostics, and insecticides, as well as the spread of the invasive mosquito species *Anopheles stephensi* (2–6).(7–9). Another major gap is the lack of vaccines that provide long-lasting immunity (10,11).

Artemisinin partial resistance (ART-R) has been linked to partial loss-of-function mutations in the *Plasmodium falciparum* Kelch13 (K13) propeller domain (4). Alarmingly, multiple ART-R mutations have emerged and are increasingly reported across the Great Rift Valley and into Southern Africa (6,12–18). World Health Organization (WHO)-validated and candidate K13 mutations such as P441L, C469Y, C469F, R561H, R622I, and A675V have been identified in these regions and appear to be increasing in frequency and spreading across borders (6,12–17). These appear to be de novo mutations that originated in Africa (6,12,15). Given the full mutational landscape that can impart ART-R has not been fully characterized, it is important to continue to monitor for the emergence of novel mutations that may impart ART-R in addition to previously validated mutations.

Artesunate-amodiaquine (ASAQ) and artemether-lumefantrine (AL) were the first artemisinin-based combination therapies (ACT) used as first-line treatment for uncomplicated malaria in Gabon, but recent policy changes in 2024 now recommend ASAQ, AL, artesunate-mefloquine (ASMQ) and dihydroartemisinin-piperaquine (DP). Sulfadoxine-pyrimethamine (SP) is used for intermittent preventive treatment in pregnancy, while severe malaria is treated with injectable artemisinin derivatives or quinine. The national malaria control program, in partnership with the Université des Sciences de la Santé, provides free malaria diagnosis at six sentinel sites and collects blood samples for molecular surveillance. However, to date, molecular drug resistance surveillance in Gabon has relied on less sensitive restriction fragment length polymorphism (RFLP) methods and was limited to a few loci analyzed by study according to a recent review (19). To date, no validated or candidate ART-R mutations have been reported in Gabon (20,21), and ACT efficacy remains high, with cure rates above 95% for AL and ASAQ (22,23).

To support malaria control, next-generation sequencing (NGS) offers a high-throughput, and detailed approach to understand parasite mutations, their origins, and their spread (24,25). Molecular surveillance also provides insights into malaria transmission dynamics, parasite relatedness, and selective pressures on the parasite population (26–28). This type of genomic surveillance has not been recently conducted in Gabon. Therefore, we conducted a pilot genomic surveillance project using samples collected between 2021 and 2023 from a peri-urban sentinel site in Libreville. We employed molecular inversion probe (MIP) capture and Illumina deep sequencing to assess antimalarial drug resistance, infection complexity, and parasite relatedness. Our analysis revealed non-synonymous mutations in K13, including one mutation that appears to be increasing in frequency over time.

## RESULTS

We sequenced 537 samples with parasitemia ≥ 500 parasites/μl (by microscopy) collected between 2021-2023 in Melen, a peri-urban sentinel site in Libreville. We successfully genotyped 465 samples to sufficient depth, defined as unique molecular identifier (UMI) counts per MIP of ≥ 5, for variant calling using DR23KE and IBC2CORE panels (29–31). We analyzed 77 samples from 2021, 169 from 2022 and 219 from 2023 for drug resistance mutations, relatedness, and complexity of infection (COI).

### Uncharacterized non-synonymous mutations in K13 BTB/POZ and propeller domains are emerging in Gabon

While no WHO validated or candidate K13 mutations were detected, we found one mutation in the BTB/POZ domain (E433D) and three uncharacterized mutations within the propeller domains (V520I, Q613H and V637I) (**Fig.1A**)(32). The most common mutation was E433D – first being seen in 2022 in one (0.6%) of 154 samples. The mutation prevalence increased to 6.3% (13/207) in 2023, the last year surveyed. The Q613H mutation was detected in 5 isolates (5/204, 2.4%) in 2023. The V520I (1/67) and V637I (1/79) only occurred in only one participant each in 2021. Many (10/21) of these propeller domain and BTB/POZ mutations were either the only or the dominant allele found within an infection and there was no evidence of multiple mutations within a haplotype (**Fig1.B**). The prevalence of the A578S, which is a wide-spread mutation not associated with ART-R, was low with 1.2% (1/81, CI) in 2021 and 1.4% (3/221) in 2023. Outside the BTB/POZ and propeller domain, ten polymorphisms were detected (**Table S1**).

**Figure 1:**
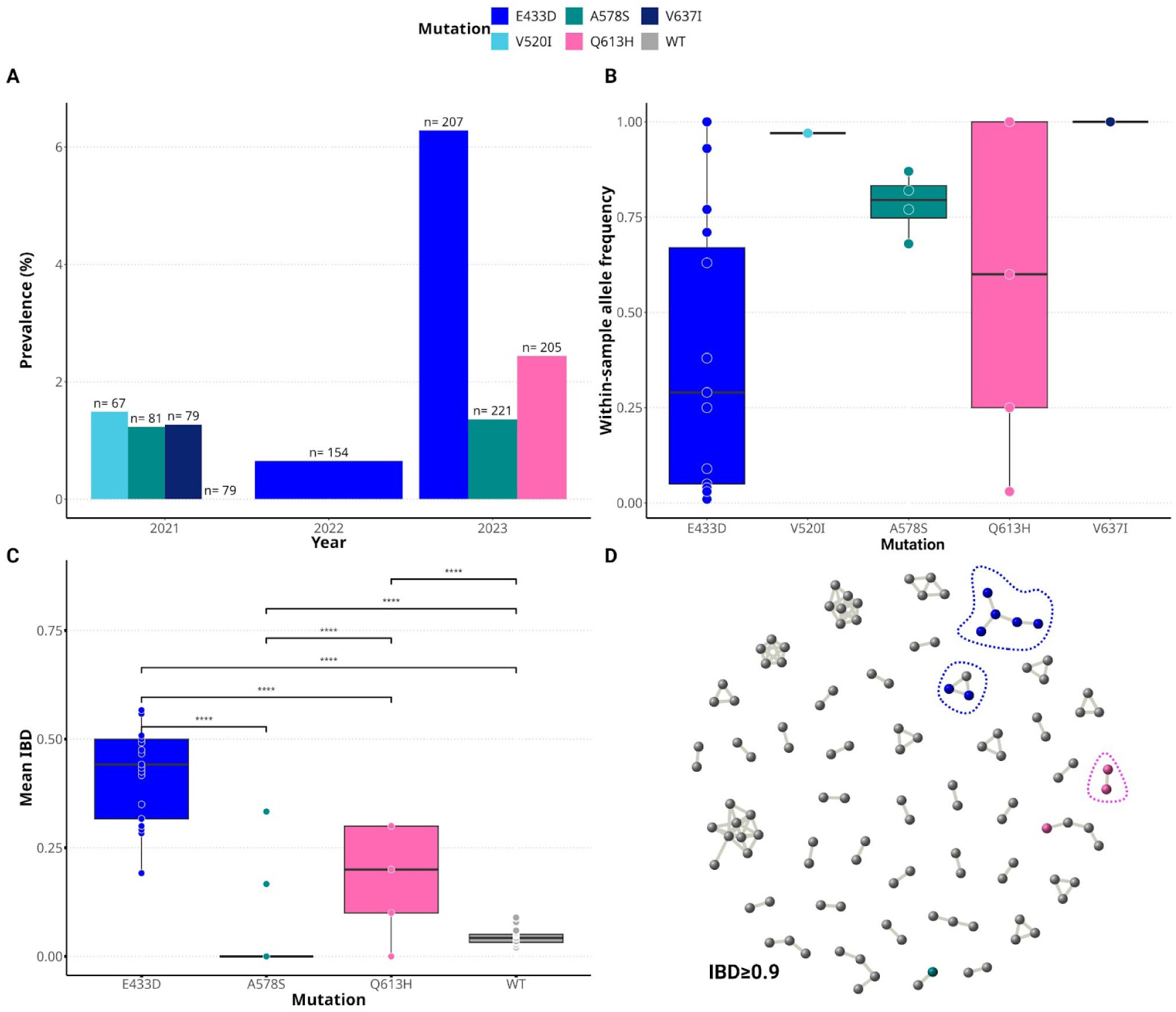
Prevalence of non-synonymous mutations detected in the *k13* gene and parasite relatedness. **A)** Prevalence of mutations by year. Sample sizes are shown on top of the barplot. **B)** Fraction of unique molecular identifiers (UMIs) supporting each mutation within-sample defined as within-sample allele frequency. The minimum total UMI count by mutation site was 17. **C)** Relatedness between isolates carrying the same mutations versus wildtype (WT) alleles based on IBD. **D)** IBD networks showing clusters of samples carrying E433D and Q613H despite at IBD≥0.90. IBD analysis was performed with all samples (n=465). Clusters of samples carrying the same K13 mutations are indicated by dotted lines.

To understand the relatedness of parasites carrying the same K13 mutation, we performed pairwise identity-by-descent (IBD) analysis between them using IBC2CORE data which targeted SNPs outside the drug resistance genes (29). The mean IBD fractions between sample pairs with E433D (mean IBD=0.42) and Q613H (mean IBD=0.2) were significantly higher (*P<0.0001,* **Fig.1C**) compared to wildtype (WT) sample pairs (mean IBD=0.04) and A578S sample pairs (mean IBD=0.03). IBD network analysis showed clustering of samples carrying E433D and Q613H at IBD>0.9, consistent with clonal chains of transmission (**Fig1.D**).

### Prevalence of most antimalarial drug resistance mutations remains stable over time

We analyzed SP resistance markers and found that dihydrofolate reductase (DHFR) mutations N51I, C59R, and S108N were fixed, with prevalence consistently above 97% over time (**Fig. 2A & B**). The I164L mutation in DHFR was not detected. Similarly, the dihydropteroate synthase (DHPS) A437G mutation was fixed (>97% prevalence across all three years). Other common DHPS mutations included S436A and K540E (**Fig. 2B**). While K540E remained stable at ∼5%, S436A increased from 7.6% (5/66) in 2021 to 18.2% (28/154) in 2022 and 23.1% (49/212) in 2023. A581G and A613S were detected only in 2022 and 2023, with prevalence below 1% (2/217) and 3% (4/156), respectively. The triple DHFR mutation combination (N51I+C59R+S108N, or IRN) was present in over 90% of samples each year (**Table I**). The quintuple mutation IRN+A437G+K540E (IRN+GE) declined from 9.7% (6/62) in 2021 to 5.3% (11/209) in 2023. No sextuple mutations were detected.

**Figure2 :**
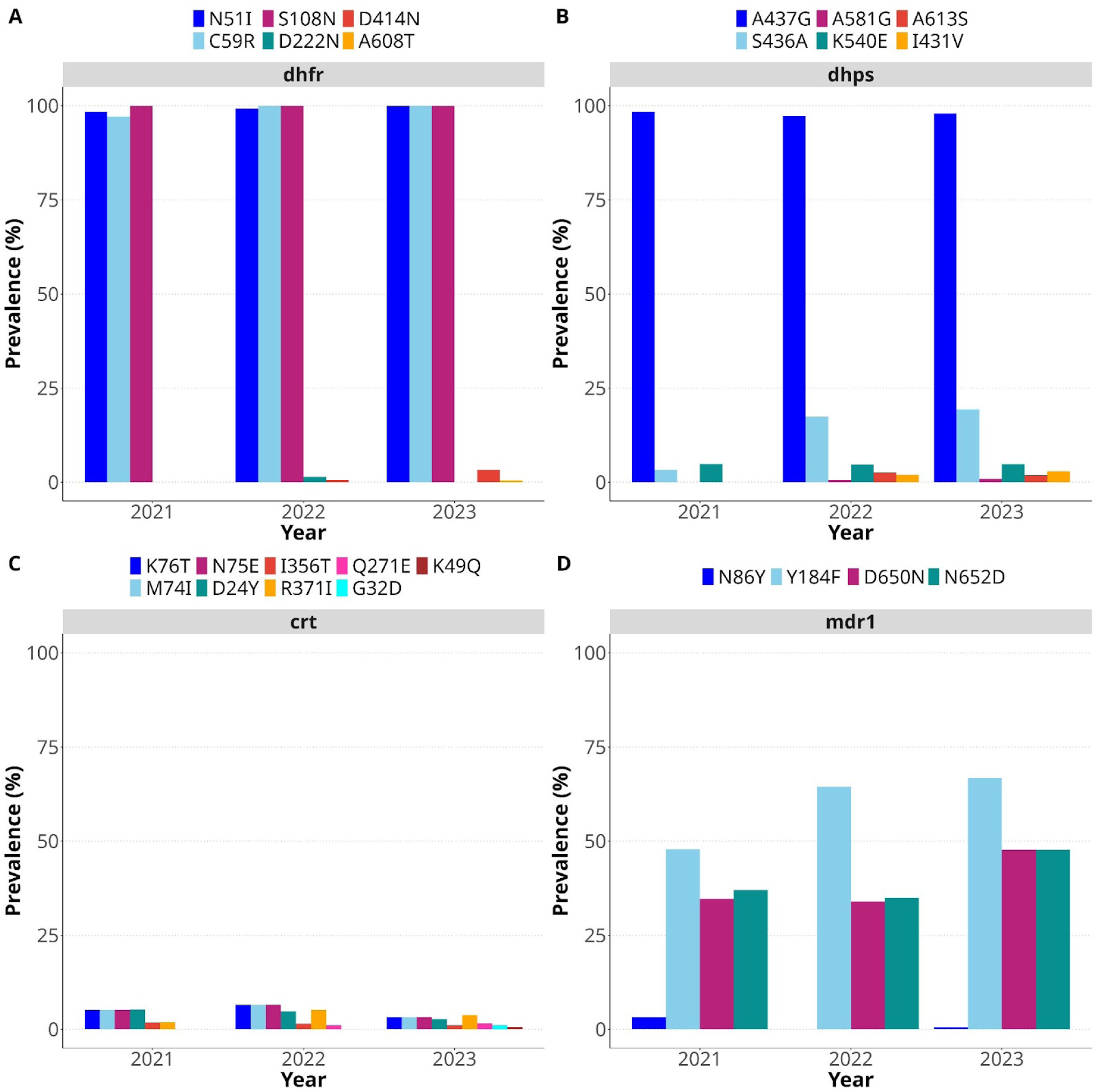
Prevalence of common antimalarial drug resistance mutations from 2021 to 2023. Prevalence of DHFR (**A**), DHPS (**B**), CRT (**C**) and MDR1 (**D**) mutations, respectively (n=465). For MDR1, only major mutations, N86Y, Y184F, D650N and N652D, are shown. The rest of mutations detected are shown in supplementary Table S2.

**Table 1:** Prevalence of DHFR triple mutations and DHFR and DHPS quintuple mutations by year.

| Haplotype | Frequency % (n/N) |  |  | Number of Mutations |
| --- | --- | --- | --- | --- |
|  | 2021 | 2022 | 2023 |  |
| IRN | 90.3 (56/62) | 92.9 (145/156) | 94.7 (198/209) | Triple |
| IRN+GE | 9.7 (6/62) | 7.1 (11/156) | 5.3 (11/209) | Quintuple |

To assess resistance to partner drugs amodiaquine and lumefantrine, we examined the chloroquine resistance transporter (CRT) and multidrug resistance 1 (MDR1) mutations, where these drugs select for mutations and wildtype, respectively. CRT mutations were rare, with the highest prevalence of the K76T mutation at 6.5% (9/139) in 2022 (**Fig. 2C**). Among MDR1 mutations, N86Y was found only in 2021 and 2023 at <3%, while Y184F increased from 52.6% (40/76) in 2021 to 68.8% (148/215) in 2023 (**Fig. 3C**). D650N and N652D remained stable between 34.6% and 47.7%. Rare MDR1 variants are listed in **Table S1**. No gene duplications were observed (**Fig. S1**).

**Figure 3:**
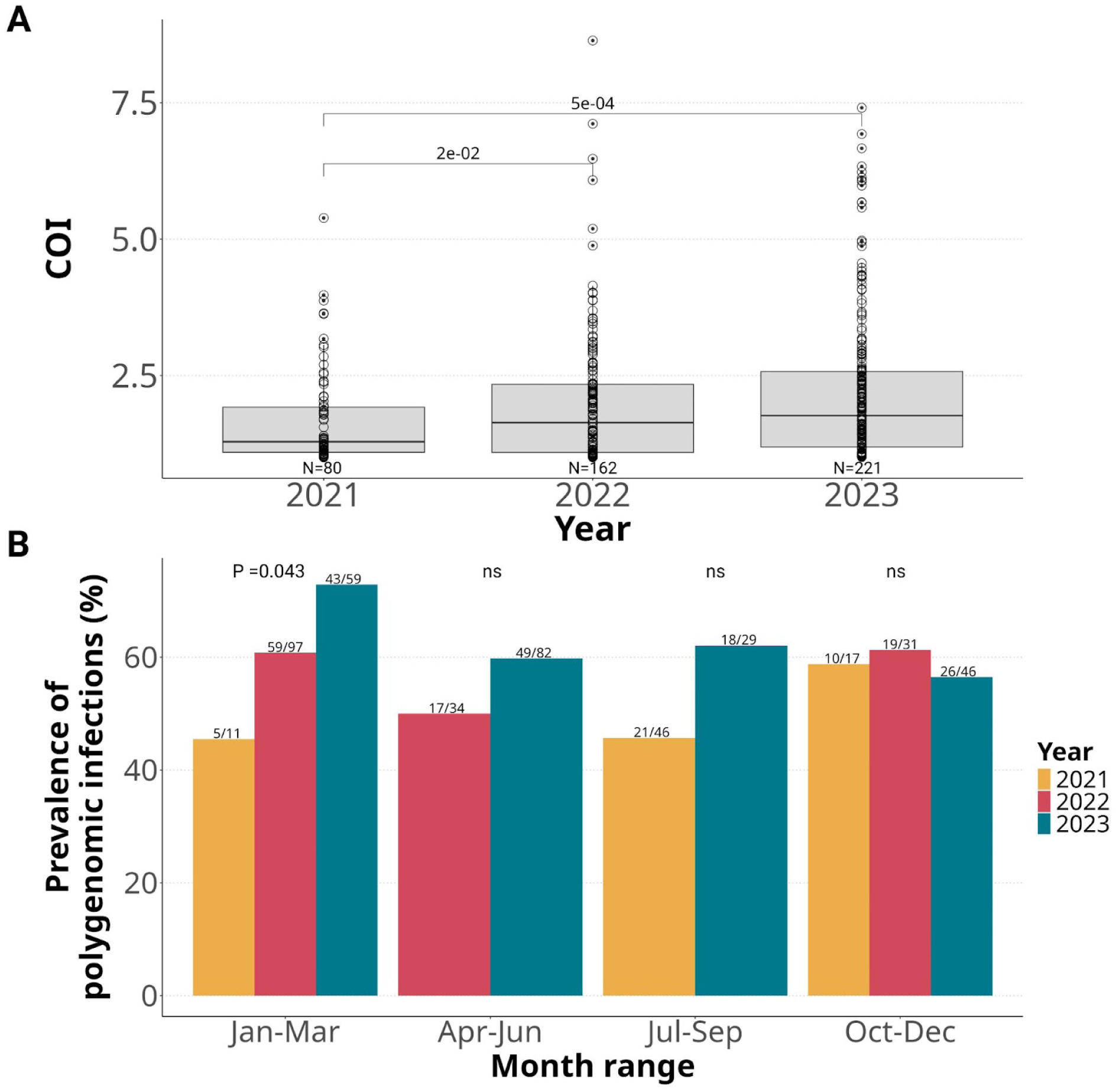
Temporal trend of complexity of infection (COI). **A)** continuous COI distribution by year. Significant pairwise comparisons of COI between years are indicated by the *P* values displayed (Wilcoxon Test). The temporal increase of COI between 2021–2023 was significant (*P<0.0009*) using a linear regression model. Dots represent samples and sample size is indicated below each box. **B**) Comparison of proportions of polygenomic infections (fraction of samples with COI>1) between 2021–2023 after every three-month period. Significance of the linear regression analysis between proportion of polygenomic infections and year is indicated on top of the bars as well as polygenomic sample count and sample size.

We also investigated additional resistance-associated loci relevant in Africa, including including putative amino acid transporter (AAT1), plasmepsin 2 and 3 (PM2 and PM3), cysteine proteinase falcipain 2a (FP2a), GTP cyclohydrolase 1 (GCH1), and coronin (CORONIN) (**Table S2**). In AAT1, the S258L mutation was nearly fixed (96.7–100%), while F313S was absent. No duplications of *pm2 or pm3*—linked to piperaquine resistance—were found. Although PM3 SNPs were rare, PM2 Q442H was frequent (86.7–90.6%). In FP2A, only Q414H was detected (59.3–66.4%). GCH1 mutations were rare, with no evidence of copy number variation. In coronin, mutations G50E, R100K, and E107V were absent, while S183G was the most common (73.3–83.3%).

### Complexity of infection varies by year

To assess recent changes in malaria transmission intensity in western Gabon, we analyzed the distribution of Complexity of Infection (COI) over time at our sentinel site (**Fig. 3**). Overall, COI showed a significant upward trend across the study period, with a strong positive correlation with time (P < 0.0009, linear regression). Although the increase from 2022 to 2023 approached significance, it still suggests a potential intensification of transmission during this period (**Fig. 3A**).

Given that malaria transmission in Gabon occurs year-round (33–35), we further investigated whether the observed rise in COI was seasonally driven. We calculated the proportion of polygenomic infections (samples with COI > 1) in quarterly intervals and compared these proportions across the three survey years. From January to September, polygenomic infections generally increased between 2021 and 2023 (**Fig. 3B**), with a statistically significant rise during January–March (P = 0.043, linear regression). However, from October to December, the proportions remained relatively stable across years.

### Genetic relatedness between samples suggests some clonal transmission of parasites from the same year and those with E433D and Q613H mutations

To assess the genetic relatedness of malaria parasites within and across years, we analyzed identity-by-descent (IBD) fractions across all 71,610 sample pairs, which provides insights into parasite interrelatedness (36). This analysis was based on 669 genome-wide SNPs generated using the IBC2CORE panel. Within-year pairwise IBD fractions—representing the proportion of the genome with shared IBD between sample pairs—increased steadily from 2021 to 2023 (**Fig. 4A**). To further explore population structure, we conducted an IBD network analysis at varying thresholds of relatedness. At IBD ≥ 0.50, the majority of isolates formed a single interconnected network, regardless of the year of collection (**Fig. 4B**). However, when the threshold was raised to IBD ≥ 0.90, clustering became more year-specific, revealing multiple groups of highly related (near-clonal) parasites, mostly from the same year (**Fig. 4C**). A few clusters spanned multiple years, including those containing E433D mutant samples.

**Figure 4:**
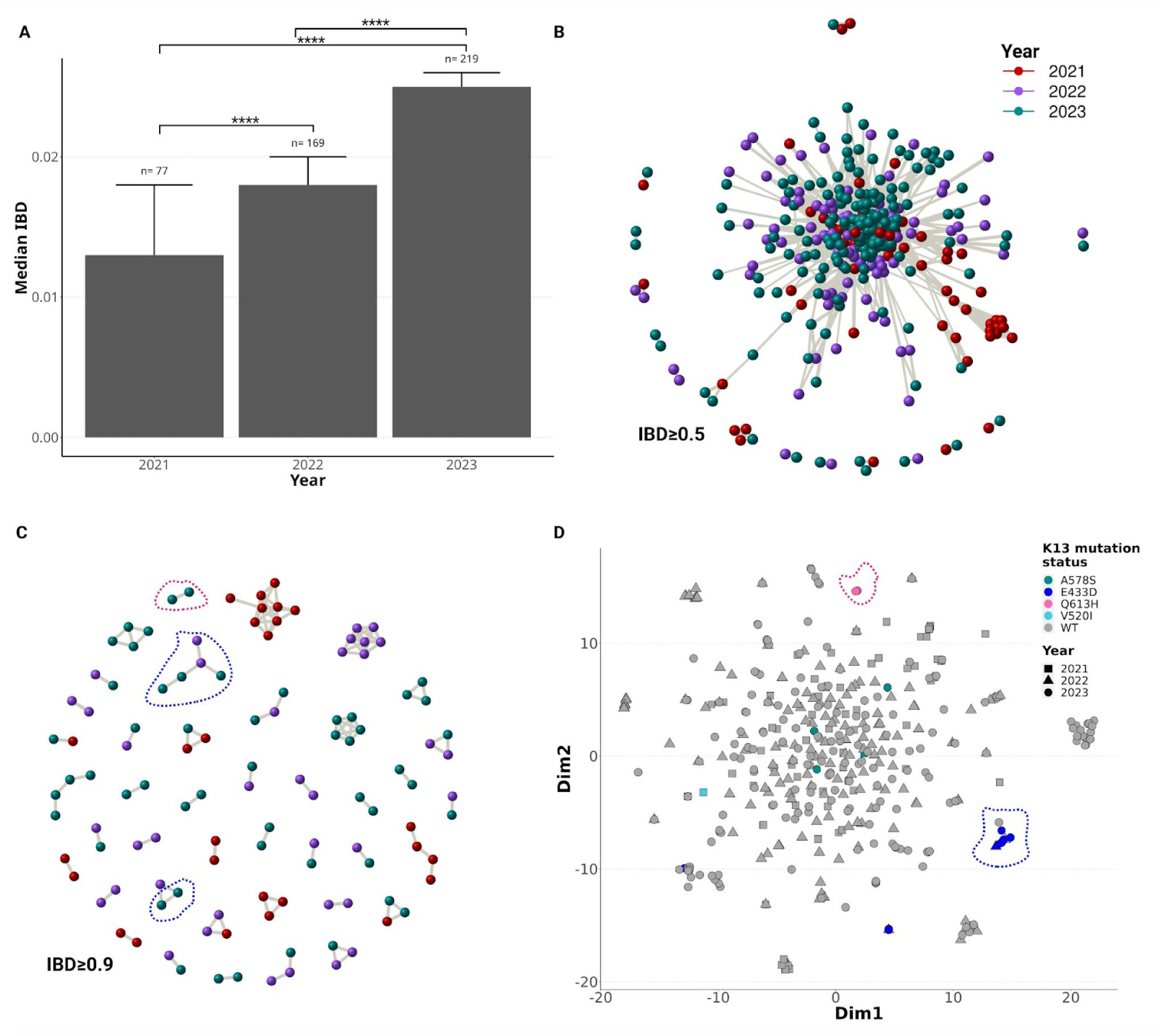
Relatedness between samples within and across years. **A)** Comparison of pairwise IBD fractions across years. IBD fractions between isolate pairs significantly over time (* indicates significance). **B)** IBD network analysis at IBD ≥0.50 showing the vast majority of samples clustering into a single network of relatives. **C)** IBD network analysis at IBD ≥95 showing samples clustering by year. Each dot represents an isolate and color codes for sample collection year. All the samples were included in IBD analysis (n=465). **D**) Population structure partially defined by sample collection year and K13 mutation (n=465). Clusters of samples carrying the same K13 mutations are circled by colored dotted lines.

We also performed t-distributed Stochastic Neighbor Embedding (t-SNE) analysis using the same SNP dataset to investigate potential clustering by year and/or K13 mutation status. While most samples did not form distinct clusters, several year-specific clusters were observed (**Fig. 4D**). Additionally, some clusters included samples from different years, such as a group of E433D mutants. Q613H isolates also formed a distinct cluster, whereas A578S mutants were dispersed among the unclustered samples (**Fig. 4D**).

## DISCUSSION

This three-year pilot genomic surveillance project in Gabon identified emerging mutations in the K13 gene, particularly E433D in the BTB/POZ domain and Q613H in the propeller domain. While these are not validated or candidate ART-R mutations, their appearance is concerning. Particularly, E433D showed a notable increase in frequency between 2022 and 2023, while Q613H appeared in the most recent year. Both showed evidence of clonal chains of transmission. These findings, along with a rising COI suggestive of increasing transmission and spreading markers MDR1 N86 and 184F associated with decreased sensitivity to lumefantrine, highlight the importance of continued monitoring and further investigation into the potential implications for malaria control in the region.

The E433D substitution appears to be a newly emerging mutation in Gabon, first detected in 2022 and rising to a prevalence of 6% by 2023. This ten-fold increase over a single year suggests strong selective pressure, potentially driven by widespread use of ACTs. The mutation was not previously reported in Gabon or neighboring countries, indicating a local emergence (19–21,37). Genetic analyses revealed that E433D was primarily found in mixed infections and showed high IBD among carriers, suggesting clonal expansion. In addition to E433D, three propeller domain mutations—V520I, Q613H, and V637I—were identified. Among these, Q613H was the most prevalent (2.4%) and may warrant close monitoring. While V520I and V637I were rare and only detected in 2021, Q613H showed signs of clonal transmission similar to E433D. Although different amino acid substitutions at positions 613 and 637 have been reported in other African countries, these specific variants are novel in Gabon (37–41).

The study site, a peri-urban area of Libreville, is characterized by mesoendemic and unstable malaria transmission, which presents a unique epidemiological setting for the emergence and spread of drug-resistant parasites (33,34). Historical data show fluctuating malaria prevalence in this region, with a notable decline from 34% in 2005 to 25% in 2011, followed by a resurgence to 31% in 2021, indicating a recent upward trend (33,34). This pattern of instability may reflect changes in vector control efforts, environmental conditions, and healthcare access over time. Such fluctuations can lead to inconsistent levels of population immunity, particularly in children and other vulnerable groups, which in turn may increase susceptibility to infection and facilitate the selection of resistant strains (42,43). The combination of unstable transmission and high drug pressure—driven by widespread ACT use and easy access to antimalarials—creates favorable conditions for resistance to emerge (12,42,43). Informal drug markets and self-medication practices may contribute to suboptimal treatment, intensifying selective pressure (44,45).

Surveillance also revealed concerning trends in resistance to partner drugs used in ACTs, as well as SP. The MDR1 N86 wild-type allele was found to be fixed in the population, a shift that likely reflects long-term selection pressure from the widespread use of AL, the most commonly used ACT in Gabon (46). In parallel, the Y184N mutation in MDR1 showed a rising prevalence, which may further indicate adaptation to lumefantrine exposure (47–49). Although the functional implications of Y184N remain under investigation, its increasing frequency suggests it may play a role in modulating drug response or compensating for fitness costs associated with other resistance mutations. In contrast, the CRT K76T mutation—associated with resistance to amodiaquine (48)—remained rare, suggesting that ASAQ is not widely used or has not exerted strong selective pressure in this setting. This supports the continued efficacy of ASAQ as an effective treatment option in Gabon. SP is primarily used in pregnant women for intermittent preventive treatment although trimethoprim and sulfamethoxazole is preferred in HIV-infected women to prevent opportunistic infections. Markers of SP resistance were widespread, with fixation of DHFR mutations (N51I, C59R, S108N) and high prevalence of DHPS A437G. Notably, the DHPS S436A mutation associated with resistance to sulfadoxine showed a rising trend from 7.6% in 2021 to 23.1% in 2023, indicating increasing SP pressure. However, key late-stage mutations associated with high-level SP resistance—such as DHFR I164L and DHPS K540E, A581G, and A613S (50–61)—remained rare or absent. This suggests that, despite the widespread presence of early SP resistance markers, SP may still retain some utility in Gabon, particularly for intermittent preventive treatment in pregnancy. These results are also in keeping with well controlled use of SP, which is rarely utilized for self-medication in Gabon.

This study was limited to a single geographic site in northwestern Gabon, which may not fully represent the genetic diversity or resistance dynamics occurring in other regions of the country or Central Africa more broadly. Sample collection was mainly disrupted by the COVID-19 pandemic, leading to relatively small sample sizes, particularly in 2021; which may limit the statistical power to detect low-frequency mutations or subtle trends over time. While identity-by-descent and population structure analyses suggested clonal transmission of key mutations such as E433D and Q613H, these findings would benefit from confirmation through whole genome sequencing to better understand the broader genomic context and evolutionary history of these variants.

In conclusion, this pilot study provides important early insights into the evolving genetic landscape of malaria parasites in Gabon. Although none of the K13 mutations identified—E433D, Q613H, V520I, and V637I—have been validated as markers of artemisinin resistance, the prevalence of E433D and Q613H with evidence of clonal transmission warrant further examination. Functional studies, such as CRISPR-Cas9–based gene editing and *in vitro* drug susceptibility assays, are needed to determine whether these mutations confer reduced artemisinin sensitivity. Together, these findings emphasize the need for expanded genomic surveillance, phenotypic validation, and integrated monitoring of both artemisinin and partner drug resistance to inform malaria control strategies in Central Africa.

## MATERIALS AND METHODS

### Study design

Surveillance was conducted at the Centre Hospitalier Régional de l’Estuaire in Melen, located in a peri-urban area 11 km from Libreville, the capital city of Gabon. This is one of six sentinel sites supported by the national malaria control program to perform malaria epidemiological surveillance and therapeutic efficacy studies. Between 2021-2023, symptomatic (≥ 37.5°C) children and adolescents from paediatric wards were screened by microscopy and persons with microscopically confirmed *P. falciparum* infection were enrolled after obtaining informed consent from guardians. Clinical and sociodemographic data were recorded. Dried blood spots (DBS) on filter paper were collected from finger prick (∼40 µL) for molecular studies. Each sample was stored in a ziplock bag containing two desiccants and kept at 4°C in a fridge before punching and shipping to Brown University for sequencing. *P. falciparum*-infected samples with at least 1,000 parasites/µL were included in the study (n=537). Ethical approval was obtained from the Scientific Committee of Comité National de Pilotage de la Riposte contre le Coronavirus-Gabon (reference: 0072/P/COPIL-CS-COVID-19). The work at Brown on deidentified samples was considered nonhuman subjects.

### Sample Processing and MIP genotyping

DNA was extracted from DBS using a Chelex-Tween protocol (30,31) before MIP capture. MIP capture and sequencing was conducted as previously described (30,31). Briefly, we combined two panels, DR23KE, targeting drug resistance genes, and IBC2CORE, capturing common SNPs across the genome for relatedness and COI analyses. This MIP pool was used to capture genomic regions of interest into circularized DNA, followed by digestion of single stranded/linearized DNA using exonucleases, to enrich captured DNA Sample barcoding, unique molecular identifier, and library preparation was conducted by PCR amplification to create final Illumina libraries that are pooled and sequenced on an Illumina Nextseq 550 at Brown University. Laboratory strains 3D7 and Dd2 were used as controls.

We used the MIPTools (version4.0) (https://github.com/bailey-lab/MIPTools) for the data analysis, including read processing and mapping and variant calling using Freebayes (version1.3.5) (Garrison & Marth, 2012). Samples with median unique molecular identifier (UMI) count < 10 across probes were repooled and resequenced to improve read depth. We used the MIPLicorn R package (https://github.com/bailey-lab/miplicorn) to process the output of the MIPTools analysis before calculating the prevalence of mutations found in drug resistance genes. Copy number of *mdr1, pfpm2* and *pfpm3* was measured by calculating read depth fold-change per MIP, based on UMI counts, relative to other non-copy number regions captured by the DR23KE panel as previously described (62).

### Complexity of infection analysis

The SNP genotypes generated with IBC2CORE MIPs was used for the discrete COI calculation using THE REAL McCOIL package (version 2.0) (63). We set the total number of Markov chain Monte Carlo and burn-in iterations to 2000 with 500, respectively. Only targeted SNPs for which the MIPs were designed with site and sample missingness of <10% and <20%, respectively, were used. The proportion of polygenomic infections, samples with discrete COI > 1 over total sample size, was calculated by year and for three month intervals. The continuous COI was estimated using the *coiaf* R package based on the same filtered SNPs (64). This COI calculation was corrected for the variation of site-level read depth.

### Identity-by-descent analysis

We used the hmmIBD package (v3.0) to compute IBD fractions between sample pairs based on SNPs obtained with IBC2CORE MIPs. The maximum number of fit iterations was set to 5 to detect recent big IBD blocks. A theoretical genotype error rate of 0.1% was adopted and clustering and visualization were performed using the *igraph* R package.

## Authors contribution

NNJM, BAKMK, JJJ, JAB and KN designed and conducted the study. KN analyzed data and interpreted results. JJJ and JAB supervised the analysis and interpretation of the data. JJJ and JAB obtained primary funding for the molecular work. MMD, MJC, MNP, KL, MMDP, NNJM and BAKMK performed sample collection and shared metadata. NNJM led the field work in Gabon. KN and AL performed DNA isolation and sequencing. KN wrote the primary draft of the manuscript. All authors contributed to the writing of the manuscript.

## Acknowledgements

The authors are grateful to Malaria Research and Reference Reagent Resource Center (MR4) for sharing laboratory strain DNA controls. The authors thank study participants and caregivers at the Melen sentinel site.

## Data Availability

Parasite sequences that support the findings of this study have been deposited in the Sequence Read Archive repository under the accession numbers SAMN50482924 to SAMN50483388 at https://www.ncbi.nlm.nih.gov/bioproject/PRJNA1302560

## References

1. World Health Organization. World malaria report 2024 [Internet]. World Health Organization; 2024 [cited 2025 May 1]. Available from: https://www.who.int/publications/i/item/9789240104440

2. World Health Organization. World malaria report 2023. World Health Organization; 2023.

3. Dondorp AM, Nosten F, Yi P, Das D, Phyo AP, Tarning J, et al. Artemisinin resistance in Plasmodium falciparum malaria. N Engl J Med. 2009;361(5):455–67.

4. Ariey F, Witkowski B, Amaratunga C, Beghain J, Langlois AC, Khim N, et al. A molecular marker of artemisinin-resistant Plasmodium falciparum malaria. Nature. 2014 Jan 2;505(7481):50–5.

5. Ashley E a., Dhorda M, Fairhurst RM, Amaratunga C, Lim P, Suon S, et al. Spread of artemisinin resistance in Plasmodium falciparum malaria. N Engl J Med. 2014;371(5):411–23.

6. Uwimana A, Legrand E, Stokes BH, Ndikumana JLM, Warsame M, Umulisa N, et al. Emergence and clonal expansion of in vitro artemisinin-resistant Plasmodium falciparum kelch13 R561H mutant parasites in Rwanda. Nat Med. 2020 Oct;26(10):1602–8.

7. Balkew M, Mumba P, Dengela D, Yohannes G, Getachew D, Yared S, et al. Geographical distribution of Anopheles stephensi in eastern Ethiopia. Parasit Vectors. 2020 Jan 20;13(1):35.

8. Seyfarth M, Khaireh BA, Abdi AA, Bouh SM, Faulde MK. Five years following first detection of Anopheles stephensi (Diptera: Culicidae) in Djibouti, Horn of Africa: populations established-malaria emerging. Parasitol Res. 2019 Mar;118(3):725–32.

9. Ranson H, Lissenden N. Insecticide Resistance in African Anopheles Mosquitoes: A Worsening Situation that Needs Urgent Action to Maintain Malaria Control. Trends Parasitol. 2016 Mar;32(3):187–96.

10. RTS,S Clinical Trials Partnership. Efficacy and safety of RTS,S/AS01 malaria vaccine with or without a booster dose in infants and children in Africa: final results of a phase 3, individually randomised, controlled trial. Lancet. 2015 Jul 4;386(9988):31–45.

11. RTSS Clinical Trials Partnership. Efficacy and Safety of the RTS,S/AS01 Malaria Vaccine during 18 Months after Vaccination: A Phase 3 Randomized, Controlled Trial in Children and Young Infants at 11 African Sites. Krishna S, editor. PLoS Med. 2014 Jul 29;11(7):e1001685.

12. Conrad MD, Asua V, Garg S, Giesbrecht D, Niaré K, Smith S, et al. Evolution of Partial Resistance to Artemisinins in Malaria Parasites in Uganda. N Engl J Med. 2023 Aug 24;389(8):722–32.

13. Young NW, Gashema P, Giesbrecht D, Munyaneza T, Maisha F, Mwebembezi F, et al. High frequency of artemisinin partial resistance mutations in the great lake region revealed through rapid pooled deep sequencing [Internet]. bioRxiv. 2024. Available from: https://www.medrxiv.org/content/10.1101/2024.04.29.24306442v1.abstract

14. Ishengoma DS, Mandara CI, Bakari C, Fola AA, Madebe RA, Seth MD, et al. Evidence of artemisinin partial resistance in North-western Tanzania: clinical and drug resistance markers study. medRxiv [Internet]. 2024 Feb 1 [cited 2024 Oct 3]; Available from: https://pubmed.ncbi.nlm.nih.gov/38352311/

15. Juliano JJ, Giesbrecht DJ, Simkin A, Fola AA, Lyimo BM, Pereus D, et al. Country wide surveillance reveals prevalent artemisinin partial resistance mutations with evidence for multiple origins and expansion of high level sulfadoxine-pyrimethamine resistance mutations in northwest Tanzania. medRxiv [Internet]. 2023 Nov 30; Available from: 10.1101/2023.11.07.23298207

16. Mihreteab S, Platon L, Berhane A, Stokes BH, Warsame M, Campagne P, et al. Increasing prevalence of artemisinin-resistant HRP2-negative malaria in Eritrea. N Engl J Med. 2023 Sep 28;389(13):1191–202.

17. Fola AA, Feleke SM, Mohammed H, Brhane BG, Hennelly CM, Assefa A, et al. Plasmodium falciparum resistant to artemisinin and diagnostics have emerged in Ethiopia. Nat Microbiol. 2023 Oct;8(10):1911–9.

18. Martin AC, Sadler JM, Simkin A, Musonda M, Katowa B, Matoba J, et al. Emergence and rising prevalence of artemisinin partial resistance marker Kelch13 P441L in a low malaria transmission setting in southern Zambia [Internet]. medRxiv. 2025 [cited 2025 May 2]. p. 2025.01.02.24319706. Available from: https://www.medrxiv.org/content/10.1101/2025.01.02.24319706v1.abstract

19. Sima-Biyang YV, Ontoua SS, Longo-Pendy NM, Mbou-Boutambe C, Makouloutou-Nzassi P, Moussadji CK, et al. Epidemiology of malaria in Gabon: A systematic review and meta-analysis from 1980 to 2023. J Infect Public Health. 2024 Jul;17(7):102459.

20. Kamau E, Campino S, Amenga-Etego L, Drury E, Ishengoma D, Johnson K, et al. K13-propeller polymorphisms in plasmodium falciparum parasites from sub-saharan Africa. J Infect Dis. 2015 Apr 15;211(8):1352–5.

21. Dinzouna-Boutamba SD, Iroungou BA, Akombi FL, Yacka-Mouele L, Moon Z, Aung JM, et al. Assessment of genetic polymorphisms associated with malaria antifolate resistance among the population of Libreville, Gabon. Malar J. 2023 Jun 14;22(1):183.

22. Adegbite BR, Edoa JR, Honkpehedji YJ, Zinsou FJ, Dejon-Agobe JC, Mbong-Ngwese M, et al. Monitoring of efficacy, tolerability and safety of artemether–lumefantrine and artesunate–amodiaquine for the treatment of uncomplicated Plasmodium falciparum malaria in Lambaréné, Gabon: an open-label clinical trial. Malar J. 2019 Dec 16;18(1):424.

23. Ndong Ngomo JM, Ondzagha Megnie GJ, Moutombi Ditombi B, Koumba Lengongo JV, M’Bondoukwé NP, Offouga CL, et al. Persistence of high in vivo efficacy and safety of artesunate-amodiaquine and artemether-lumefantrine as the first- and second-line treatments for uncomplicated Plasmodium falciparum malaria 10 years after their implementation in Gabon. Acta Parasitol. 2019 Dec;64(4):898–902.

24. Volkman SK, Neafsey DE, Schaffner SF, Park DJ, Wirth DF. Harnessing genomics and genome biology to understand malaria biology. Nat Rev Genet. 2012 Apr 12;13(5):315–28.

25. Golumbeanu M, Edi CAV, Hetzel MW, Koepfli C, Nsanzabana C. Bridging the gap from molecular surveillance to programmatic decisions for malaria control and elimination. Am J Trop Med Hyg. 2025 Jan 7;112(1_Suppl):35–47.

26. Schaffner SF, Badiane A, Khorgade A, Ndiop M, Gomis J, Wong W, et al. Malaria surveillance reveals parasite relatedness, signatures of selection, and correlates of transmission across Senegal. Nat Commun. 2023 Nov 10;14(1):7268.

27. Wesolowski A, Taylor AR, Chang HH, Verity R, Tessema S, Bailey JA, et al. Mapping malaria by combining parasite genomic and epidemiologic data. BMC Med. 2018 Oct 18;16(1):190.

28. Tessema S, Wesolowski A, Chen A, Murphy M, Wilheim J, Mupiri AR, et al. Using parasite genetic and human mobility data to infer local and cross-border malaria connectivity in Southern Africa. Elife [Internet]. 2019 Apr 2 [cited 2025 Jul 3];8. Available from: 10.7554/eLife.43510

29. Niaré K, Crudale R, Fola AA, Wernsman Young N, Asua V, Conrad M, et al. Highly multiplex molecular inversion probe panel in Plasmodium falciparum targeting common SNPs approximates whole genome sequencing assessments for selection and relatedness [Internet]. medRxiv. 2025. p. 2025.03.07.25323597. Available from: https://www.medrxiv.org/content/10.1101/2025.03.07.25323597v1

30. Verity R, Aydemir O, Brazeau NF, Watson OJ, Hathaway NJ, Mwandagalirwa MK, et al. The impact of antimalarial resistance on the genetic structure of Plasmodium falciparum in the DRC. Nat Commun. 2020 Apr 30;11(1):2107.

31. Aydemir O, Janko M, Hathaway NJ, Verity R, Mwandagalirwa MK, Tshefu AK, et al. Drug-resistance and population structure of Plasmodium falciparum across the Democratic Republic of Congo using high-throughput molecular inversion probes. J Infect Dis. 2018 Aug 14;218(6):946–55.

32. Malaria: Artemisinin partial resistance [Internet]. [cited 2025 Jun 18]. Available from: https://www.who.int/news-room/questions-and-answers/item/artemisinin-resistance

33. Mawili-Mboumba DP, Bouyou Akotet MK, Kendjo E, Nzamba J, Medang MO, Mbina JRM, et al. Increase in malaria prevalence and age of at risk population in different areas of Gabon. Malar J. 2013 Jan 2;12(1):3.

34. Moutombi Ditombi BC, Pongui Ngondza B, Manomba Boulingui C, Mbang Nguema OA, Ndong Ngomo JM, M’Bondoukwé NP, et al. Malaria and COVID-19 prevalence in a population of febrile children and adolescents living in Libreville. S Afr J Infect Dis. 2022 Oct 26;37(1):459.

35. Lendongo-Wombo JB, Oyegue-Liabagui SL, Biteghe-Bi-Essone JC, Ngoungou EB, Lekana-Douki JB. Epidémiology of malaria from 2019 to 2021 in the southeastern city of Franceville, Gabon. BMC Public Health. 2022 Dec 10;22(1):2313.

36. Taylor AR, Schaffner SF, Cerqueira GC, Nkhoma SC, Anderson TJC, Sriprawat K, et al. Quantifying connectivity between local Plasmodium falciparum malaria parasite populations using identity by descent. Didelot X, editor. PLoS Genet. 2017 Oct 27;13(10):e1007065.

37. Milong Melong CS, Peloewetse E, Russo G, Tamgue O, Tchoumbougnang F, Paganotti GM. An overview of artemisinin-resistant malaria and associated Pfk13 gene mutations in Central Africa. Parasitol Res. 2024 Jul 18;123(7):277.

38. Huang F, Yan H, Xue JB, Cui YW, Zhou SS, Xia ZG, et al. Molecular surveillance of pfcrt, pfmdr1 and pfk13-propeller mutations in Plasmodium falciparum isolates imported from Africa to China. Malar J. 2021 Feb 6;20(1):73.

39. Ménard D, Khim N, Beghain J, Adegnika AA, Shafiul-Alam M, Amodu O, et al. A worldwide map of Plasmodium falciparum K13-propeller polymorphisms. N Engl J Med. 2016 Jun 23;374(25):2453–64.

40. Taylor SM, Parobek CM, DeConti DK, Kayentao K, Coulibaly SO, Greenwood BM, et al. Absence of putative artemisinin resistance mutations among Plasmodium falciparum in Sub-Saharan Africa: a molecular epidemiologic study. J Infect Dis. 2015 Mar 1;211(5):680–8.

41. Schmedes SE, Patel D, Dhal S, Kelley J, Svigel SS, Dimbu PR, et al. Plasmodium falciparum kelch 13 Mutations, 9 Countries in Africa, 2014-2018. Emerg Infect Dis. 2021 Jul;27(7):1902–8.

42. Kamya MR, Nankabirwa JI, Arinaitwe E, Rek J, Zedi M, Maiteki-Sebuguzi C, et al. Dramatic resurgence of malaria after 7 years of intensive vector control interventions in Eastern Uganda. PLOS Glob Public Health. 2024 Aug 29;4(8):e0003254.

43. Namuganga JF, Epstein A, Nankabirwa JI, Mpimbaza A, Kiggundu M, Sserwanga A, et al. The impact of stopping and starting indoor residual spraying on malaria burden in Uganda. Nat Commun. 2021 May 11;12(1):2635.

44. Takyi A, Carrara VI, Dahal P, Przybylska M, Harriss E, Insaidoo G, et al. Characterisation of populations at risk of sub-optimal dosing of artemisinin-based combination therapy in Africa. PLOS Glob Public Health. 2023 Dec 1;3(12):e0002059.

45. White NJ. Antimalarial drug resistance. J Clin Invest. 2004 Apr 15;113(8):1084–92.

46. Ndong Ngomo JM, Mawili-Mboumba DP, M’Bondoukwé NP, Ditombi BM, Koumba Lengongo JV, Batchy Ognagosso FB, et al. Drug resistance molecular markers of Plasmodium falciparum and severity of malaria in febrile children in the sentinel site for malaria surveillance of Melen in Gabon: Additional data from the Plasmodium diversity network African network. Trop Med Infect Dis. 2023 Mar 23;8(4):184.

47. Sisowath C, Petersen I, Veiga MI, Mårtensson A, Premji Z, Björkman A, et al. In vivo selection of Plasmodium falciparum parasites carrying the chloroquine-susceptible pfcrt K76 allele after treatment with artemether-lumefantrine in Africa. J Infect Dis. 2009 Mar 1;199(5):750–7.

48. Venkatesan M, Gadalla NB, Stepniewska K, Dahal P, Nsanzabana C, Moriera C, et al. Polymorphisms in Plasmodium falciparum chloroquine resistance transporter and multidrug resistance 1 genes: parasite risk factors that affect treatment outcomes for P. falciparum malaria after artemether-lumefantrine and artesunate-amodiaquine. Am J Trop Med Hyg. 2014 Oct;91(4):833–43.

49. Okell LC, Reiter LM, Ebbe LS, Baraka V, Bisanzio D, Watson OJ, et al. Emerging implications of policies on malaria treatment: genetic changes in the Pfmdr-1 gene affecting susceptibility to artemether-lumefantrine and artesunate-amodiaquine in Africa. BMJ Glob Health. 2018 Oct 19;3(5):e000999.

50. Wang P, Read M, Sims PF, Hyde JE. Sulfadoxine resistance in the human malaria parasite Plasmodium falciparum is determined by mutations in dihydropteroate synthetase and an additional factor associated with folate utilization. Mol Microbiol. 1997 Mar;23(5):979–86.

51. Omar SA, Adagu IS, Warhurst DC. Can pretreatment screening for dhps and dhfr point mutations in Plasmodium falciparum infections be used to predict sulfadoxine-pyrimethamine treatment failure? Trans R Soc Trop Med Hyg. 2001 May;95(3):315–9.

52. Desai M, Gutman J, Taylor SM, Wiegand RE, Khairallah C, Kayentao K, et al. Impact of sulfadoxine-pyrimethamine resistance on effectiveness of intermittent preventive therapy for malaria in pregnancy at clearing infections and preventing low birth weight. Clin Infect Dis. 2016 Feb 1;62(3):323–33.

53. Esu E, Tacoli C, Gai P, Berens-Riha N, Pritsch M, Loescher T, et al. Prevalence of the Pfdhfr and Pfdhps mutations among asymptomatic pregnant women in Southeast Nigeria. Parasitol Res. 2018 Mar;117(3):801–7.

54. Tchuenkam PVK, Ngum LN, Ali IM, Chedjou JPK, Nji AM, Netongo PM, et al. Plasmodium falciparum dhps and dhfr markers of resistance to sulfadoxine–pyrimethamine five years (2016–2020) after the implementation of seasonal malaria chemoprevention in Cameroon. Wellcome Open Res. 2024 Jun 20;9:323.

55. Djontu JC, Baina MT, Mbama Ntabi JD, Lissom A, Umuhoza DM, Assioro Doulamo NV, et al. Profile of molecular markers of Sulfadoxine-Pyrimethamine-resistant Plasmodium falciparum in individuals living in southern area of Brazzaville, Republic of Congo. Int J Parasitol Drugs Drug Resist. 2024 Dec;26(100569):100569.

56. Nana RRD, Bayengue SSB, Mogtomo MLK, Ngane ARN, Singh V. Anti-folate quintuple mutations in Plasmodium falciparum asymptomatic infections in Yaoundé, Cameroon. Parasitol Int. 2023 Feb;92(102657):102657.

57. Nkemngo FN, Raissa LW, Nguete DN, Ndo C, Fru-Cho J, Njiokou F, et al. Geographical emergence of sulfadoxine-pyrimethamine drug resistance-associated P. falciparum and P. malariae alleles in co-existing Anopheles mosquito and asymptomatic human populations across Cameroon. Antimicrob Agents Chemother. 2023 Dec 14;67(12):e0058823.

58. L’Episcopia M, Doderer-Lang C, Perrotti E, Priuli GB, Cavallari S, Guidetti C, et al. Polymorphism analysis of drug resistance markers in Plasmodium falciparum isolates from Benin. Acta Trop. 2023 Sep;245(106975):106975.

59. Apinjoh TO, Mugri RN, Miotto O, Chi HF, Tata RB, Anchang-Kimbi JK, et al. Molecular markers for artemisinin and partner drug resistance in natural Plasmodium falciparum populations following increased insecticide treated net coverage along the slope of mount Cameroon: cross-sectional study. Infect Dis Poverty. 2017 Nov 6;6(1):136.

60. Mbacham WF, Evehe MSB, Netongo PM, Ateh IA, Mimche PN, Ajua A, et al. Efficacy of amodiaquine, sulphadoxine-pyrimethamine and their combination for the treatment of uncomplicated Plasmodium falciparum malaria in children in Cameroon at the time of policy change to artemisinin-based combination therapy. Malar J. 2010 Jan 27;9(1):34.

61. Guémas E, Coppée R, Ménard S, du Manoir M, Nsango S, Makaba Mvumbi D, et al. Evolution and spread of Plasmodium falciparum mutations associated with resistance to sulfadoxine-pyrimethamine in central Africa: a cross-sectional study. Lancet Microbe. 2023 Dec;4(12):e983–93.

62. Popkin-Hall ZR, Niaré K, Crudale R, Simkin A, Fola AA, Sanchez JF, et al. High-Throughput Genotyping of Plasmodium vivax in the Peruvian Amazon via Molecular Inversion Probes. medRxiv [Internet]. 2024 Jun 28; Available from: 10.1101/2024.06.27.24309599

63. Chang HH, Worby CJ, Yeka A, Nankabirwa J, Kamya MR, Staedke SG, et al. THE REAL McCOIL: A method for the concurrent estimation of the complexity of infection and SNP allele frequency for malaria parasites. PLoS Comput Biol. 2017 Jan;13(1):e1005348.

64. Paschalidis A, Watson OJ, Aydemir O, Verity R, Bailey JA. coiaf: directly estimating complexity of infection with allele frequencies [Internet]. bioRxiv. 2022 [cited 2022 Sep 7]. p. 2022.05.26.493561. Available from: https://www.biorxiv.org/content/10.1101/2022.05.26.493561v1

