## Supplemental materials for "Clonal Transmission of Emerging Novel *Plasmodium falciparum* Kelch13 Mutations and Increasing Complexity of Infection in Libreville, Gabon, 2021-2023"

*equal senior author contribution

✝corresponding authors:

Karamoko Niare, PhD

### Supplementary Tables

#### Table S1: Prevalence of non-propeller domain mutations of PfK13

| Gene name | Mutation | Mutation count | Sample size | Prevalence (%) | Year |
|---|---|---|---|---|---|
| k13 | G112E | 2 | 163 | 1.23 | 2023 |
| k13 | K123N | 1 | 163 | 0.61 | 2023 |
| k13 | H136N | 10 | 129 | 7.75 | 2023 |
| k13 | T149S | 1 | 135 | 0.74 | 2022 |
| k13 | T149S | 4 | 162 | 2.47 | 2023 |
| k13 | K189N | 14 | 146 | 9.59 | 2022 |
| k13 | K189N | 8 | 188 | 4.26 | 2023 |
| k13 | K189P | 1 | 154 | 0.65 | 2023 |
| k13 | K189T | 26 | 61 | 42.62 | 2021 |
| k13 | K189T | 72 | 143 | 50.35 | 2022 |
| k13 | K189T | 104 | 185 | 56.22 | 2023 |
| k13 | N217H | 3 | 202 | 1.49 | 2023 |
| k13 | R255K | 2 | 71 | 2.82 | 2021 |
| k13 | R255K | 8 | 153 | 5.23 | 2022 |
| k13 | R255K | 6 | 210 | 2.86 | 2023 |
| k13 | L258M | 2 | 71 | 2.82 | 2021 |
| k13 | L258M | 1 | 153 | 0.65 | 2022 |

#### Table S2: Other mutations detected in protein involved in drug resistance

| Gene name | Mutation | Mutation count | Sample size | Prevalence (%) | Year |
|---|---|---|---|---|---|
| PfCYTB | A11T | 1 | 149 | 0.67 | 2022 |
| PfCYTB | A11T | 3 | 202 | 1.49 | 2023 |
| PfCYTB | W154L | 2 | 217 | 0.92 | 2023 |
| PfCYTB | I179S | 1 | 226 | 0.44 | 2023 |
| PfCYTB | M308I | 6 | 148 | 4.05 | 2022 |
| PfCYTB | M308I | 3 | 195 | 1.54 | 2023 |
| PfCYTB | R364K | 6 | 162 | 3.7 | 2022 |

| | | | | | |
|---|---|---|---|---|---|
| PfCYTB | R364K | 10 | 224 | 4.46 | 2023 |
| PfMDR1 | E7Q | 3 | 216 | 1.39 | 2023 |
| PfMDR1 | N12T | 1 | 75 | 1.33 | 2021 |
| PfMDR1 | N12T | 3 | 156 | 1.92 | 2022 |
| PfMDR1 | N12T | 3 | 216 | 1.39 | 2023 |
| PfMDR1 | K34R | 2 | 145 | 1.38 | 2022 |
| PfMDR1 | K34R | 3 | 198 | 1.52 | 2023 |
| PfMDR1 | A487E | 1 | 55 | 1.82 | 2021 |
| PfMDR1 | A487E | 2 | 144 | 1.39 | 2022 |
| PfMDR1 | A487E | 1 | 175 | 0.57 | 2023 |
| PfMDR1 | Y492F | 1 | 144 | 0.69 | 2022 |
| PfMDR1 | N504K | 2 | 142 | 1.41 | 2022 |
| PfMDR1 | N504K | 4 | 174 | 2.3 | 2023 |
| PfMDR1 | T526A | 1 | 71 | 1.41 | 2021 |
| PfMDR1 | T526A | 1 | 156 | 0.64 | 2022 |
| PfMDR1 | T526A | 1 | 215 | 0.47 | 2023 |
| PfMDR1 | R640T | 5 | 188 | 2.66 | 2023 |
| PfMDR1 | D642N | 1 | 29 | 3.45 | 2021 |
| PfMDR1 | D642N | 2 | 68 | 2.94 | 2022 |
| PfMDR1 | D642N | 1 | 90 | 1.11 | 2023 |
| PfMDR1 | N649D | 2 | 29 | 6.9 | 2021 |
| PfMDR1 | N649D | 3 | 67 | 4.48 | 2022 |
| PfMDR1 | N649D | 1 | 91 | 1.1 | 2023 |
| PfMDR1 | D651N | 4 | 31 | 12.9 | 2021 |
| PfMDR1 | D651N | 6 | 68 | 8.82 | 2022 |
| PfMDR1 | D651N | 2 | 91 | 2.2 | 2023 |
| PfMDR1 | A750E | 1 | 149 | 0.67 | 2022 |
| PfMDR1 | I832L | 1 | 62 | 1.61 | 2021 |
| PfMDR1 | I832L | 4 | 146 | 2.74 | 2022 |
| PfMDR1 | I832L | 4 | 201 | 1.99 | 2023 |
| PfMDR1 | L836F | 2 | 203 | 0.99 | 2023 |
| PfMDR1 | I839V | 1 | 146 | 0.68 | 2022 |
| PfMDR1 | F938Y | 1 | 150 | 0.67 | 2022 |
| PfMDR1 | F938Y | 2 | 201 | 1 | 2023 |
| PfMDR1 | G968A | 6 | 74 | 8.11 | 2021 |

| | | | | | |
|---|---|---|---|---|---|
| PfMDR1 | G968A | 6 | 158 | 3.8 | 2022 |
| PfMDR1 | G968A | 2 | 214 | 0.93 | 2023 |
| PfMDR1 | S973N | 1 | 74 | 1.35 | 2021 |
| PfMDR1 | S973N | 11 | 158 | 6.96 | 2022 |
| PfMDR1 | S973N | 13 | 212 | 6.13 | 2023 |
| PfMDR1 | G1113A | 1 | 141 | 0.71 | 2022 |
| PF3D7_1115700 | Q414E | 32 | 54 | 59.26 | 2021 |
| PF3D7_1115700 | Q414E | 93 | 140 | 66.43 | 2022 |
| PF3D7_1115700 | Q414E | 104 | 174 | 59.77 | 2023 |
| PF3D7_1224000 | H17D | 2 | 151 | 1.32 | 2022 |
| PF3D7_1224000 | H17D | 1 | 191 | 0.52 | 2023 |
| PF3D7_1224000 | E38K | 2 | 151 | 1.32 | 2022 |
| PF3D7_1224000 | E38K | 5 | 188 | 2.66 | 2023 |
| PF3D7_1224000 | K47N | 1 | 72 | 1.39 | 2021 |
| PF3D7_1224000 | D72N | 1 | 150 | 0.67 | 2022 |
| PF3D7_1224000 | N88Y | 2 | 74 | 2.7 | 2021 |
| PF3D7_1224000 | N88Y | 1 | 156 | 0.64 | 2022 |
| PF3D7_1224000 | N88Y | 1 | 216 | 0.46 | 2023 |
| PF3D7_1224000 | L107S | 1 | 64 | 1.56 | 2021 |
| PF3D7_1224000 | G108C | 2 | 63 | 3.17 | 2021 |
| PF3D7_1224000 | G108C | 4 | 201 | 1.99 | 2023 |
| PF3D7_1224000 | S109N | 1 | 147 | 0.68 | 2022 |
| PF3D7_1224000 | S109N | 1 | 202 | 0.5 | 2023 |
| PF3D7_1224000 | I163T | 2 | 142 | 1.41 | 2022 |
| PF3D7_1224000 | I163T | 1 | 191 | 0.52 | 2023 |
| PF3D7_1224000 | R230K | 6 | 59 | 10.17 | 2021 |
| PF3D7_1224000 | R230K | 12 | 147 | 8.16 | 2022 |
| PF3D7_1224000 | R230K | 19 | 195 | 9.74 | 2023 |
| PF3D7_1224000 | Y232F | 11 | 195 | 5.64 | 2023 |
| PF3D7_1251200 | A28T | 1 | 196 | 0.51 | 2023 |
| PF3D7_1251200 | V62M | 1 | 65 | 1.54 | 2021 |
| PF3D7_1251200 | V62M | 8 | 155 | 5.16 | 2022 |
| PF3D7_1251200 | V62M | 6 | 207 | 2.9 | 2023 |
| PF3D7_1251200 | P76S | 9 | 64 | 14.06 | 2021 |
| PF3D7_1251200 | P76S | 26 | 153 | 16.99 | 2022 |

| | | | | | |
|---|---|---|---|---|---|
| PF3D7_1251200 | P76S | 45 | 207 | 21.74 | 2023 |
| PF3D7_1251200 | S183G | 62 | 74 | 83.78 | 2021 |
| PF3D7_1251200 | S183G | 110 | 150 | 73.33 | 2022 |
| PF3D7_1251200 | S183G | 167 | 206 | 81.07 | 2023 |
| PF3D7_1251200 | I375V | 1 | 210 | 0.48 | 2023 |
| PF3D7_1251200 | L412R | 2 | 204 | 0.98 | 2023 |
| PF3D7_1251200 | V424I | 13 | 73 | 17.81 | 2021 |
| PF3D7_1251200 | V424I | 24 | 157 | 15.29 | 2022 |
| PF3D7_1251200 | V424I | 32 | 202 | 15.84 | 2023 |
| PF3D7_1251200 | F434L | 7 | 73 | 9.59 | 2021 |
| PF3D7_1251200 | F434L | 8 | 156 | 5.13 | 2022 |
| PF3D7_1251200 | F434L | 7 | 201 | 3.48 | 2023 |
| PF3D7_1251200 | R435K | 1 | 154 | 0.65 | 2022 |
| PF3D7_1251200 | R435K | 4 | 199 | 2.01 | 2023 |
| PF3D7_1251200 | R435T | 1 | 199 | 0.5 | 2023 |
| PF3D7_1251200 | G449W | 1 | 150 | 0.67 | 2022 |
| PF3D7_1251200 | D458N | 1 | 63 | 1.59 | 2021 |
| PF3D7_1251200 | D458N | 5 | 150 | 3.33 | 2022 |
| PF3D7_1251200 | D458N | 4 | 197 | 2.03 | 2023 |
| PF3D7_1251200 | P509T | 1 | 152 | 0.66 | 2022 |
| PF3D7_1251200 | D528N | 1 | 79 | 1.27 | 2021 |
| PF3D7_1251200 | D528N | 1 | 167 | 0.6 | 2022 |
| PF3D7_1251200 | D528N | 1 | 221 | 0.45 | 2023 |
| PF3D7_1251200 | Q532R | 1 | 165 | 0.61 | 2022 |
| PF3D7_1251200 | Q532R | 8 | 215 | 3.72 | 2023 |
| PF3D7_1251200 | D547N | 9 | 215 | 4.19 | 2023 |
| PfAAT1 | G160S | 4 | 148 | 2.7 | 2022 |
| PfAAT1 | S258L | 60 | 62 | 96.77 | 2021 |
| PfAAT1 | S258L | 153 | 153 | 100 | 2022 |
| PfAAT1 | S258L | 202 | 204 | 99.02 | 2023 |
| PfPM2 | I15M | 1 | 59 | 1.69 | 2021 |
| PfPM2 | M51I | 1 | 142 | 0.7 | 2022 |
| PfPM2 | M51I | 1 | 191 | 0.52 | 2023 |
| PfPM2 | L80F | 2 | 148 | 1.35 | 2022 |
| PfPM2 | T81I | 1 | 148 | 0.68 | 2022 |

| | | | | | |
|---|---|---|---|---|---|
| PfPM2 | L117F | 1 | 46 | 2.17 | 2021 |
| PfPM2 | L117F | 3 | 138 | 2.17 | 2022 |
| PfPM2 | L117F | 2 | 162 | 1.23 | 2023 |
| PfPM2 | V133A | 1 | 203 | 0.49 | 2023 |
| PfPM2 | T154I | 1 | 59 | 1.69 | 2021 |
| PfPM2 | T154I | 1 | 191 | 0.52 | 2023 |
| PfPM2 | I257V | 2 | 188 | 1.06 | 2023 |
| PfPM2 | V284L | 3 | 214 | 1.4 | 2023 |
| PfPM2 | T289I | 2 | 154 | 1.3 | 2022 |
| PfPM2 | T289I | 2 | 215 | 0.93 | 2023 |
| PfPM2 | L321V | 2 | 149 | 1.34 | 2022 |
| PfPM2 | L321V | 2 | 202 | 0.99 | 2023 |
| PfPM2 | A323V | 1 | 76 | 1.32 | 2021 |
| PfPM2 | A323V | 4 | 219 | 1.83 | 2023 |
| PfPM2 | G388R | 2 | 68 | 2.94 | 2021 |
| PfPM2 | G388R | 13 | 151 | 8.61 | 2022 |
| PfPM2 | G388R | 10 | 207 | 4.83 | 2023 |
| PfPM2 | I413V | 1 | 151 | 0.66 | 2022 |
| PfPM2 | I413V | 5 | 203 | 2.46 | 2023 |
| PfPM2 | P421S | 8 | 150 | 5.33 | 2022 |
| PfPM2 | P421S | 10 | 202 | 4.95 | 2023 |
| PfPM2 | Q442H | 58 | 64 | 90.62 | 2021 |
| PfPM2 | Q442H | 130 | 150 | 86.67 | 2022 |
| PfPM2 | Q442H | 180 | 202 | 89.11 | 2023 |
| PfPM3 | R67P | 1 | 148 | 0.68 | 2022 |
| PfPM3 | K151N | 1 | 151 | 0.66 | 2022 |
| PfPM3 | K151N | 1 | 211 | 0.47 | 2023 |
| PfPM3 | V165I | 1 | 151 | 0.66 | 2022 |
| PfPM3 | P222T | 1 | 171 | 0.58 | 2023 |
| PfPM3 | V276I | 2 | 56 | 3.57 | 2021 |
| PfPM3 | P304S | 1 | 150 | 0.67 | 2022 |
| PfPM3 | V327I | 2 | 154 | 1.3 | 2022 |
| PfPM3 | V327I | 1 | 212 | 0.47 | 2023 |
| PfPM3 | L397I | 1 | 22 | 4.55 | 2021 |
| PfPM3 | L397I | 1 | 93 | 1.08 | 2022 |

| | | | | | |
|---|---|---|---|---|---|
| PfPM3 | L397I | 1 | 102 | 0.98 | 2023 |

**Supplementary Figure**

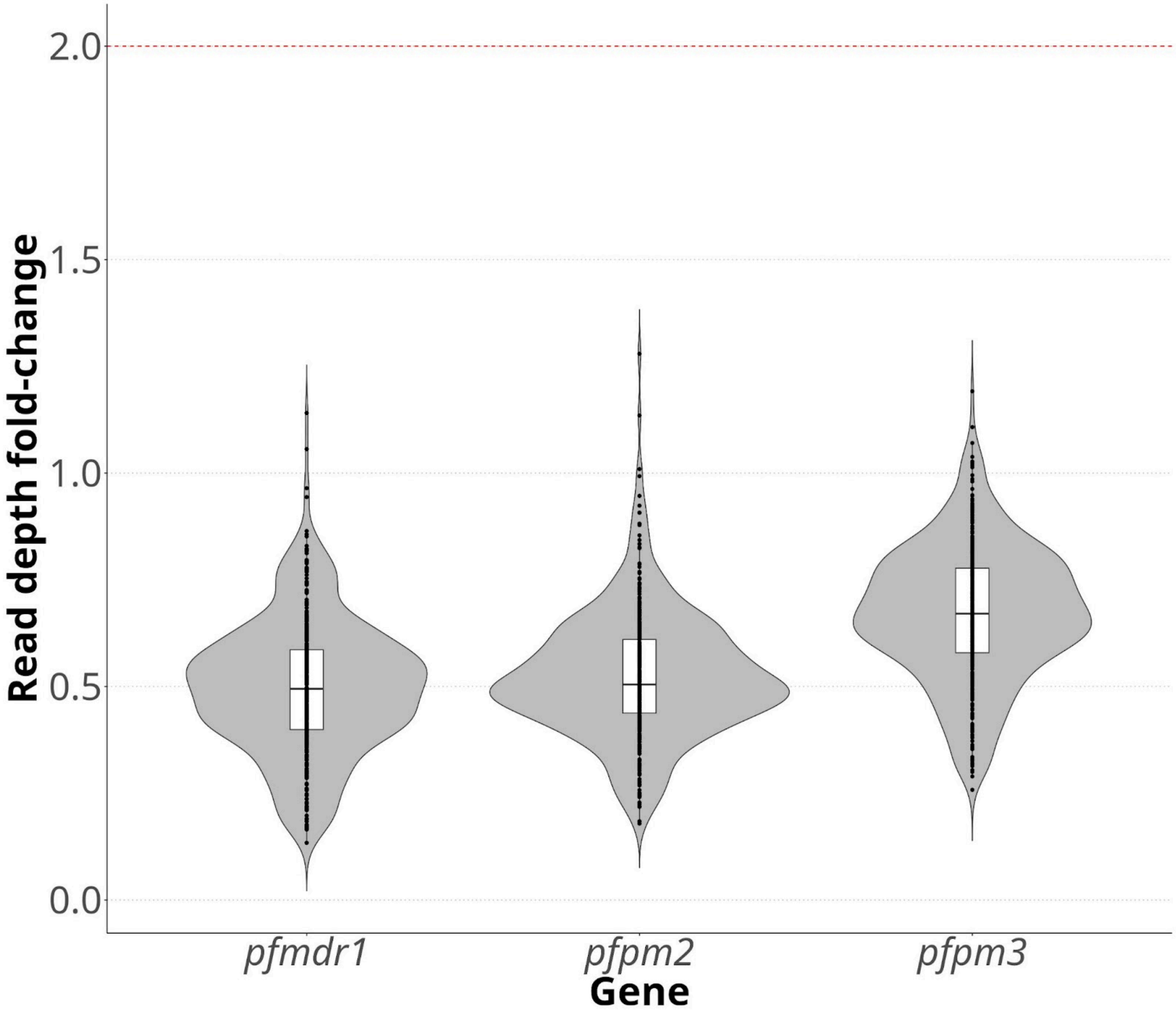


**Figure S1: Lack of number  number variation of *pfmdr1, Pfpm2 and Pfm3* gene**. Y-axis represents read depth fold-change per MIP of gene of interest relative to single copy region captured after z-score normalization.
